# CD-Tron: Leveraging Large Clinical Language Model for Early Detection of Cognitive Decline from Electronic Health Records

**DOI:** 10.1101/2024.10.31.24316386

**Authors:** Hao Guan, John Novoa-Laurentiev, Li Zhou

## Abstract

**Background:** Early detection of cognitive decline during the preclinical stage of Alzheimer’s disease and related dementias (AD/ADRD) is crucial for timely intervention and treatment. Clinical notes in the electronic health record contain valuable information that can aid in the early identification of cognitive decline. In this study, we utilize advanced large clinical language models, fine-tuned on clinical notes, to improve the early detection of cognitive decline.

**Methods:** We collected clinical notes from 2,166 patients spanning the 4 years preceding their initial mild cognitive impairment (MCI) diagnosis from the Enterprise Data Warehouse of Mass General Brigham. To train the model, we developed CD-Tron, built upon a large clinical language model that was finetuned using 4,949 expert-labeled note sections. For evaluation, the trained model was applied to 1,996 independent note sections to assess its performance on real-world unstructured clinical data. Additionally, we used explainable AI techniques, specifically SHAP values (SHapley Additive exPlanations), to interpret the model’s predictions and provide insight into the most influential features. Error analysis was also facilitated to further analyze the model’s prediction.

**Results:** CD-Tron significantly outperforms baseline models, achieving notable improvements in precision, recall, and AUC metrics for detecting cognitive decline (CD). Tested on many real-world clinical notes, CD-Tron demonstrated high sensitivity with only one false negative, crucial for clinical applications prioritizing early and accurate CD detection. SHAP-based interpretability analysis highlighted key textual features contributing to model predictions, supporting transparency and clinician understanding.

**Conclusion:** CD-Tron offers a novel approach to early cognitive decline detection by applying large clinical language models to free-text EHR data. Pretrained on real-world clinical notes, it accurately identifies early cognitive decline and integrates SHAP for interpretability, enhancing transparency in predictions.

## 1. Introduction

Alzheimer’s disease and related dementias (AD/ADRD) rank among the top 10 leading causes of death in the United States [1, 2]. It is projected that by 2060, nearly 14 million people will be living with AD/ADRD [3, 4]. AD/ADRD is a neurodegenerative disorder characterized by a gradual decline in memory and other cognitive functions. The disease typically progresses through three stages: preclinical, mild cognitive impairment (MCI), and dementia [5, 6], as shown in Fig. 1. In recent years, early detection of cognitive decline (CD), including subject cognitive decline (SCD), has gained attention as a critical stage in the continuum of cognitive aging and potential progression to AD/ADRD. SCD, characterized by self-reported memory or cognitive concerns without measurable impairment on standard tests, may serve as an early indicator of future decline, including progression to MCI or AD/ADRD. Identifying cognitive changes early allows for risk stratification, lifestyle and pharmacological interventions, and enrollment in clinical trials targeting disease-modifying treatments. Moreover, early detection supports healthcare providers in optimizing patient management, facilitating discussions about long-term care, and addressing modifiable risk factors to potentially slow progression [7, 8].

**Figure 1:**
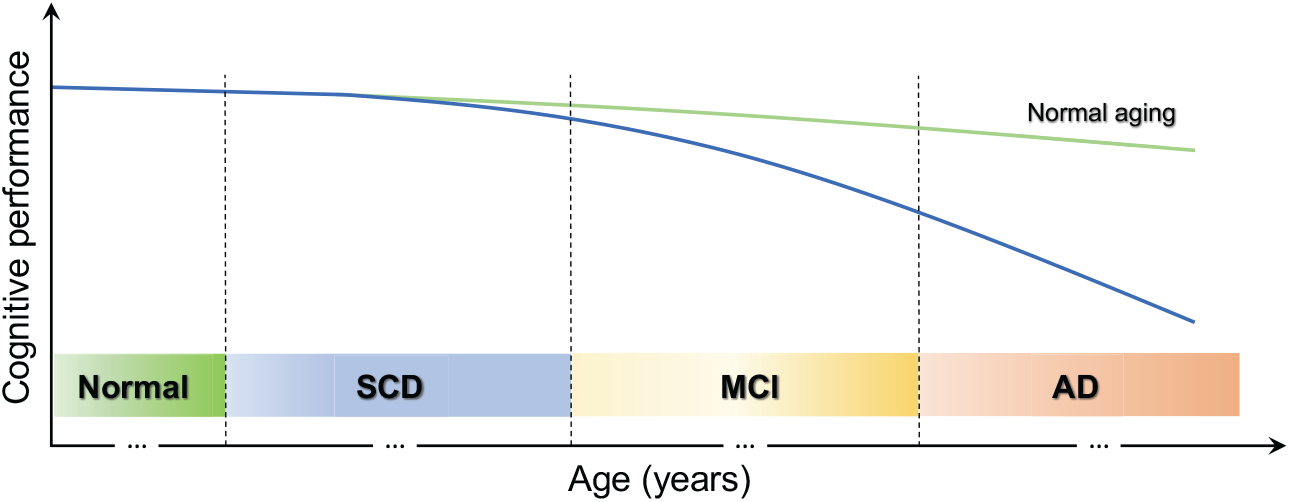
Conceptual illustration of the stages in the Alzheimer’s disease continuum.

In recent decades, various methods utilizing different data modalities, such as MRI, PET scans, cerebrospinal fluid (CSF) analysis, and genetic testing, have been developed for detecting cognitive decline [9, 10, 11, 12]. However, these methods are often invasive, expensive, and resource-intensive, which can limit their feasibility for large-scale screening. In contrast, electronic health records (EHRs) offer several advantages for cognitive decline detection [13, 14]. EHRs are cost-effective and readily accessible, making them a more practical option for widespread screening compared to high-cost imaging and laboratory-based tests. Additionally, EHRs contain extensive clinical notes that can capture subtle cognitive symptoms that may be overlooked by other methods, enabling earlier detection and intervention. Thus we focus on using the unstructured EHR data, *i.e*., clinical notes for our study.

Natural Language Processing (NLP) has emerged as a powerful tool for extracting meaningful insights from EHRs clinical notes [15, 16]. NLP facilitates the automatic analysis of large volumes of text, enabling the identification of patterns, relationships, and trends that are often difficult to detect manually [17, 18]. In recent years, transformer-based models like BERT [19] and its domain-specific variants such as ClinicalBERT [20], BioBERT [21] and MedBERT [22], have significantly enhanced the ability to extract relevant information for clinical decision-making [23, 24, 25]. Some prior studies have applied these techniques for AD or MCI-to-AD conversion prediction [26, 27, 28]. However, these models have several limitations in clinical applications. They are relatively smaller models compared to some of the more recent large-scale models, which may limit their ability to comprehend specialized medical terminology and the overall context of clinical documents. Additionally, even with fine-tuning on small medical datasets, they lack the depth of clinical knowledge needed for complex healthcare tasks. These limitations highlight the need for more advanced clinical language models in our study.

To overcome these challenges, recent developments in large language models (LLMs) have achieved great breakthrough [29, 30, 31], and also introduced more sophisticated architectures tailored for the medical domain [32, 33, 34]. In a previous study in our group, we have already tried general-domain large language models (*e.g*., GPT and Llama) [35]. Despite this potential, there have been relatively few studies leveraging large clinical language models **specifically** designed for the early detection of cognitive decline in the preclinical stage of AD/ADRD using clinical notes. Thus this work focuses on pre-trained models on medical records. Recently, models like GatorTron [36] and NYUTron [37] represent a significant leap forward, addressing the limitations of the above-mentioned models. These LLMs are trained on vastly larger and more diverse clinical datasets, encompassing a broad range of medical and general knowledge, which enhances their ability to handle biomedical related tasks. Our study aims to address this gap by demonstrating the potential of large clinical language models for efficient detection of cognitive decline. This approach opens the door to more scalable and effective screening tools for brain health.

Based on the above motivations, we propose an AI model, CD-Tron, built upon a large clinical language model for detecting cognitive decline using clinical notes. Our approach leverages the power of a state-of-the-art clinical language model to capture the complex clinical knowledge and nuanced semantics within these notes. To further enhance the interpretability of the model, we incorporate explainable AI techniques using SHAP values (SHapley Additive exPlanations), offering valuable insights into the factors influencing the model’s predictions.

### Statement of Significance

The significance of this paper can be summarized in Table 1. More specifically, this paper makes the following contributions.

**Table 1:**
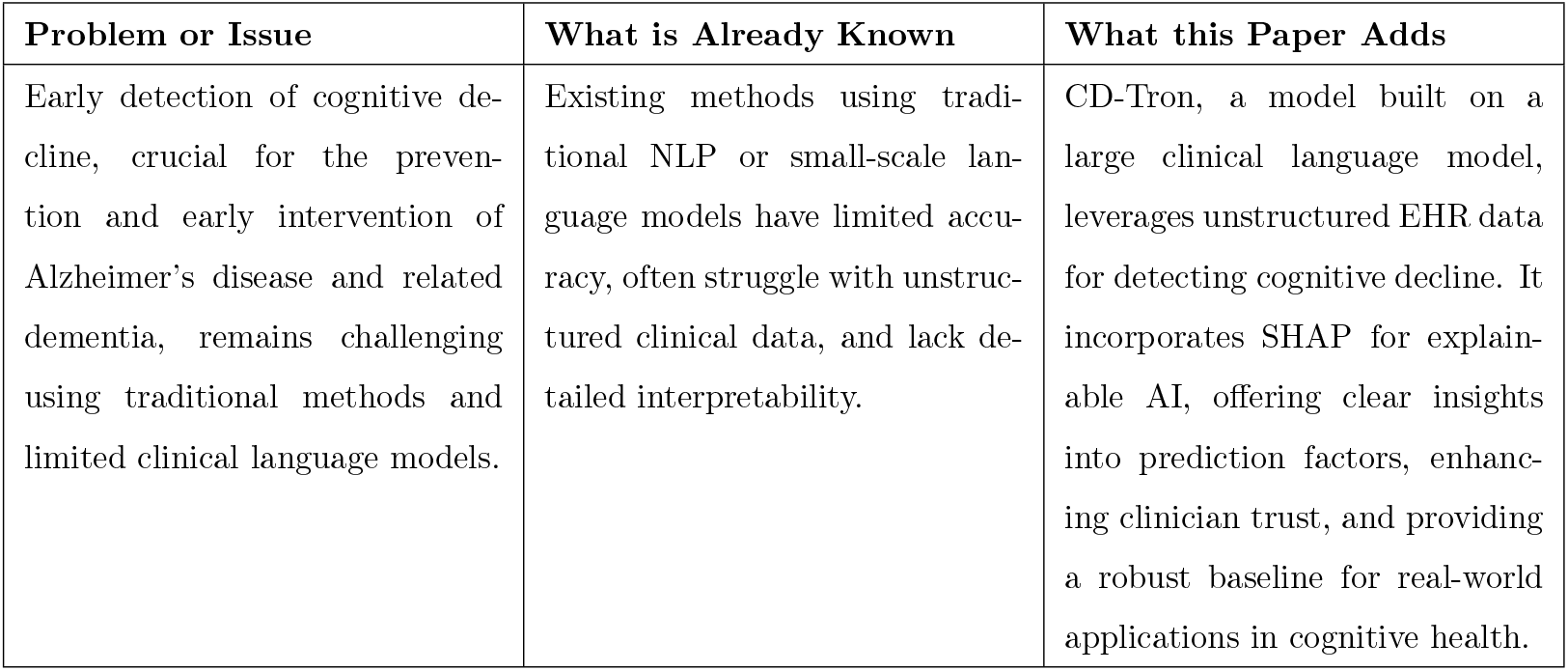
Summary of Significance.

| Problem or Issue | What is Already Known | What this Paper Adds |
| --- | --- | --- |
| Early detection of cognitive decline, crucial for the prevention and early intervention of Alzheimer’s disease and related dementia, remains challenging using traditional methods and limited clinical language models. | Existing methods using traditional NLP or small-scale language models have limited accuracy, often struggle with unstructured clinical data, and lack detailed interpretability. | CD-Tron, a model built on a large clinical language model, leverages unstructured EHR data for detecting cognitive decline. It incorporates SHAP for explainable AI, offering clear insights into prediction factors, enhancing clinician trust, and providing a robust baseline for real-world applications in cognitive health. |

1) **Integration of a Large Clinical Language Model:** Our work is one of the first to adapt large clinical language model, specifically for the early detection of cognitive decline using free-text clinical notes. This represents a significant step forward in leveraging LLMs for identifying individuals with cognitive decline, including potential preclinical stage of AD/ADRD.
2) **Explainability in Clinical Context:** By applying SHAP to interpret the model’s predictions, we introduce a novel way to gain insights into how large clinical language models make decisions. This approach not only enhances transparency but also provides actionable information that clinicians can use to understand and trust the model’s outputs.
2) **Real-World Data Application:** Our study applies these advanced techniques to a real-world dataset of free-text EHR notes, demonstrating the practical utility of our model in clinical settings.

## 2. Materials

### 2.1. Definition of Cognitive Decline

This study aims to identify patients at any stage of cognitive decline, ranging from SCD to MCI to dementia. We adopt the definition of cognitive decline from Wang *et al*. [16]. Cognitive decline was identified based on the presence of documented cognitive concerns, symptoms (*e.g*., memory loss), diagnoses (*e.g*., MCI), cognitive assessments (*e.g*., Mini-Cog), and cognitive-related therapies (*e.g*., cognitive-linguistic therapy). SCD represents an early stage, characterized by self-reported concerns such as memory loss or forgetfulness from patients or caregivers [38]. These self-reported concerns reflect perceived cognitive decline but do not constitute a formal diagnosis. Our analysis focused on progressive cognitive decline likely lead to or align with MCI. Cases of stable or improving cognitive function, transient impairment (*e.g*., temporary memory loss due to medication such as codeine), or reversible deficits (*e.g*., post-surgical or post-injury cognitive changes) were classified as negative for cognitive decline. Note sections were also labeled as negative if they contained broad or uncertain indications of cognitive decline.

### 2.2. Data Collection

This study was conducted using data from Mass General Brigham (MGB), the largest healthcare system serving the Greater Boston area. Clinical notes for patients were retrieved from MGB’s Enterprise Data Warehouse. We specifically extracted clinical notes from the four years preceding each patients initial MCI diagnosis in 2019 to focus on the early stages of cognitive decline. The clinical notes analyzed in this study were authored by a diverse group of healthcare providers, including primary care physicians, specialists, and other clinical staff. Additionally, the dataset encompasses a wide range of healthcare settings, spanning outpatient visits with specialists and inpatient care. The data usage has been approved by the Institutional Review Board (IRB) of MGB (Number: 2022P002987, 2022P002772), and all of them were deidentified before our model development.

### 2.3. Data Processing

The clinical notes were divided into sections using the NLP tool, *i.e*., Medical Text Extraction, Reasoning, and Mapping System (MTERMS) [39]. Given the variability in the length and content of clinical notes, these sections can range from highly relevant to irrelevant in terms of cognitive assessment. The clinical notes used in this study were annotated to identify sections associated with cognitive decline. To ensure the quality and clinical relevance of the annotations, a rigorous process was designed in collaboration with clinical experts and researchers specializing in cognitive decline.

### 2.4. Training and Test Datasets

We first developed a labeled dataset (Dataset I) for model development. As the mentions of CDs in notes are sparse, to increase the positive case density for effective model learning, we applied a list of expert-curated keywords to filter for sections that likely contain indications of cognitive decline. Once we identified a list of sections that included the keywords, we randomized the sections by patient and section name to account for an uneven distribution of sections among patients and section types. Dataset I contained 4,949 sections among which a total of 1,453 were labeled as cognitive decline.

To evaluate model performance on note sections independent of keyword filtering, we created a second labeled dataset (Dataset II) to evaluate the model trained on Dataset I. Dataset II consisted of 1,996 randomly selected sections from the full note corpus, excluding the sections in Dataset I. This dataset was used to assess model generalizability to unfiltered note sections, reflecting a real-world clinical scenario where mentions of CD in notes are naturally sparse. This setup allows us to evaluate the models ability to generalize beyond keyword-driven patterns and perform well in practical deployment settings. Despite the difference in positive case ratios, we ensured robust evaluation by employing stratified evaluation metrics such as Receiver Operating Characteristic Area Under the Curve (ROC-AUC), and Area under the Precision-Recall Curve (PR-AUC), which are less affected by class imbalance than accuracy-based metrics.

### 2.5. Data Annotation

Annotation criteria were carefully defined to guide the labeling process. Sections were labeled as positive if they explicitly mentioned cognitive concerns, symptoms such as memory loss or confusion, results of formal cognitive assessments (*e.g*., Mini-Cog), or diagnoses indicative of cognitive decline or MCI. Conversely, sections were labeled as negative if they referred to transient or reversible cognitive impairments, such as those caused by surgery, injury, or medication, or if they were deemed irrelevant or ambiguous with respect to cognitive decline.

The annotation process began with an initial keyword-based search to identify potentially relevant sections in the clinical notes. Each section was then independently reviewed and labeled by three trained annotators. Discrepancies in labeling were resolved through a consensus-driven process, ensuring alignment with the predefined criteria. The annotation team comprised a multidisciplinary group of researchers and clinicians, combining expertise in natural language processing and cognitive health. This collaborative approach ensured the annotations were both scientifically robust and clinically meaningful.

## 3. Methods

### 3.1. Overview

The overall pipeline of our framework is illustrated in Fig. 2. Our proposed framework, CD-Tron, was designed to detect early cognitive decline using clinical notes. All the notes were firstly segmented into different sections which were fed into to the AI model. A large pre-trained clinical language model was leveraged to extract the sematic features of each section. Finally, a binary classification network output the prediction of cognitive decline status (CD or non-CD). To enhance the interpretability of the proposed model, an explainable AI technique, *i.e*., SHAP was integrated into the framework. The SHAP values can provide interpretability for the model’s predictions.

**Figure 2:**
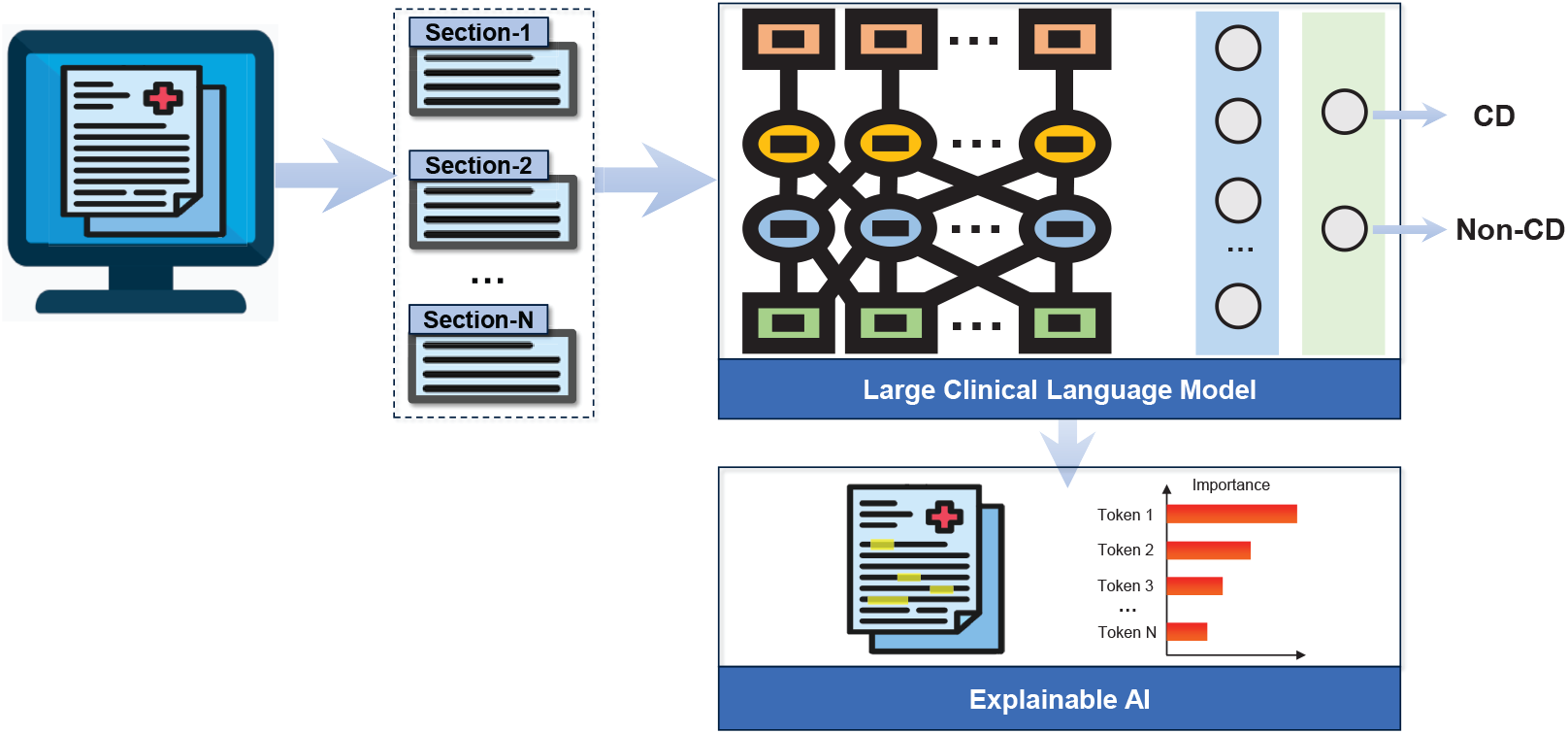
Illustration of the framework and workflow of the proposed CD-Tron based on large clinical language model for cognitive decline detection from EHRs.

### 3.2. Model Structure

The overall structure of the proposed CD-Tron model is shows in Fig. 2. The CD-Tron model was built upon GatorTron which is a large clinical language model trained on extensive biomedical and clinical text corpora. While general-domain LLMs such as GPT-4 and Gemini exceed 100 billion parameters, GatorTron stands as one of the largest biomedical-domain LLMs, scaling up to billions of parameters, far surpassing prior clinical transformer models such as ClinicalBERT with millions of parameters. As originally introduced in its foundational study [36], GatorTron represents a significant advancement in biomedical NLP, demonstrating the scalability and impact of LLMs within the clinical domain. The Transformer [40] served as the core structure of the model. The pre-training allowed the model to capture complex clinical knowledge and nuances present in the unstructured notes. We trained the model for the specific task of cognitive decline detection by feeding clinical note sections into the model and applying a classification head to output a binary prediction (CD or non-CD). To ensure interpretability, SHAP values were employed to explain model decisions, allowing the identification of the most influential tokens that contribute to each prediction.

### 3.3. Training

The CD-Tron model was trained on a dataset of clinical notes collected from patients at MGB. During training, the model processed clinical note sections as input and learned to predict whether the patient has cognitive decline or not. During training, an initial learning rate of 2e-5 was used, controlled by the AdamW optimizer. This learning rate allowed for gradual adjustments to the model weights without drastic changes.

The model was trained on a standard supervised classification task with binary cross-entropy loss as the objective function. A linear learning rate scheduler with warm-up was employed to ensure smooth convergence by slowly increasing the learning rate at the beginning of training and gradually decreasing it later. We set the maximum input length of clinical note sections to 512 tokens, enabling the model to handle lengthy text inputs common in clinical notes. We found that reducing the input length can negatively impact performance. The model was trained for 2 epochs, with each epoch involving the complete pass of the training data. This was found to provide sufficient convergence without overfitting. The proposed framework was implemented by the PyTorch and HuggingFace Transformer library [41].

### 3.4. Inference and Explainability

Inference was performed by applying the trained CD-Tron model to unseen clinical notes from the test set. For each note, the model predicted whether the patient was likely to have cognitive decline or not. To provide transparency in decision-making, we employed SHAP values [42, 43] to interpret the models outputs.

SHAP values allowed us to identify which sections or tokens in the clinical notes contributed most to the model’s decision. By assigning importance scores to each feature (word or token), SHAP offered a detailed breakdown of the factors influencing the prediction. This interpretability is particularly important in the clinical context, where understanding the rationale behind a decision is critical for clinician trust and model adoption. Visualizing these SHAP values enhanced the overall transparency of the CD-Tron model, allowing clinicians to better understand why the model predicts CD for a particular patient, fostering confidence in its predictions for real-world clinical applications.

In LLMs, another option for interpretation is using prompting to generate detailed explanations in sentence form. However, while prompts are commonly used by generative language models to create narrative explanations based on input text, they are not ideally suited for the structured interpretation required in classification tasks. Instead, we chose SHAP for its ability to precisely quantify the contribution of each input feature (word or token) to the model’s decision. SHAP values provide a clear, feature-level breakdown of the factors influencing the prediction, which is crucial in clinical contexts where understanding the specific reasons behind a decision is essential for clinician trust and model adoption. Additionally, prompt-based explanations are more limited in their application, particularly within LLMs, whereas SHAP is a model-agnostic tool. This means SHAP can be used to explain predictions from any machine learning model, significantly enhancing the flexibility and applicability of our approach.

While attention mechanisms are often considered useful for interpretability, recent studies [44, 45] have shown that attention weights do not always correlate with actual feature importance. Attention distributions primarily capture token dependencies rather than the direct influence of individual features on the final model prediction. In this study, we opted for SHAP-based analysis, as SHAP values provide a more direct measure of how specific input components contribute to the model’s decision. This approach ensures a more clinically meaningful and transparent interpretation of CD-Tron’s predictions. Therefore, we did not further explore attention-based explanations, as they may not reliably reflect the factors influencing the model’s decision-making in cognitive decline detection.

### 3.5. Contributions Summary

While CD-Tron builds upon the GatorTron model, it introduces several key adaptations to address the specific task of cognitive decline detection from clinical notes. GatorTron, like other transformer-based large language models (LLMs), was pre-trained using a self-supervised learning objective such as masked language modeling, it was not specifically designed or optimized for downstream domain-specific tasks. As such, its performance may be suboptimal when applied directly to such tasks without task-specific fine-tuning.

In this study, we aim to demonstrate how existing clinical LLMs can be effectively adapted for specialized clinical tasks by developing CD-Tron, a system specifically tailored for the early detection of cognitive decline from clinical notes. Importantly, CD-Tron is more than a reapplication of GatorTron. In particular, CD-Tron differs from GatorTron in several key ways: (1) Task-Specific Adaptation: CD-Tron includes domain-relevant preprocessing and expert-annotated data that focus specifically on cognitive decline, a task not originally considered in the development of GatorTron; (2) Fine-Tuning: We fine-tuned GatorTron on our custom-labeled dataset, optimizing the model to detect subtle cognitive signals in real-world clinical notes; and (3) the integration of SHAP values to provide token-level interpretability for predictions. The addition of interpretability is critical for clinical trust and transparency and was not addressed in the original GatorTron work. Our interpretable outputs support clinicians in understanding why a prediction was made, enhancing the models potential for real-world adoption. The above modifications form the core of the CD-Tron system and distinguish it from the original GatorTron model.

## 4. Experiment

### 4.1. Experimental Setup

#### 4.1.1. Evaluation Metrics

The task in our study was to identify if a clinical section was relevant to cognitive decline or not. A section that represented cognitive decline was regarded as the positive case (with label **1**), while the others were negative cases (with label **0**). It was a binary classification problem. For performance evaluation, we used four metrics, *i.e*., Precision, Recall, F1-score, ROC-AUC, and PR-AUC. Higher values for each evaluation metric indicated better performance.

#### 4.1.2. Competing Methods

##### Logistic Regression

We represented each section using Term Frequency-Inverse Document Frequency (TF-IDF) based on n-grams (n=1, 2). These feature vectors were then utilized to train a logistic regression classifier, which predicted whether a section indicates cognitive decline.

##### Random Forest

We used the TF-IDF features of the sections of clinical notes to train a Random Forest classifier for the task of cognitive decline detection. The key hyperparameter, *i.e*., number of trees in the forest, was set to 100.

##### Support Vector Machine (SVM)

TF-IDF features of the sections were used to train an SVM classifier (with a linear kernel) for cognitive decline detection.

##### BERT

We used the vanilla BERT for the CD detection task. The BERT model was first fine-tuned on the training set, and then applied to the test set. A fully-connected layer was used as the classification head for CD vs. non-CD classification on the sections.

##### BioBERT

The BioBERT model was pre-trained on biomedical literature (*e.g*., scientific papers from PubMed). We fine-tuned it on the training set, and then applied to the test set. A fully-connected layer was added as the classifier for CD vs. non-CD classification on the sections.

##### ClinicalBERT

The ClinicalBERT was pre-trained on the MIMIC III dataset which contained clinical notes, including discharge summaries, progress notes, etc. A fully-connected layer was built for the identification of CD on the sections.

In our experiments with these deep learning-based methods, each model was run five times, and the average and standard deviation of the results were reported.

### 4.2. Result

The training set consisted of 4,949 sections from 1,969 patients (1,046 [53.1%] female; mean age 76.0 *±* 13.3 years), while the test set included 1,996 sections from 1,161 patients (619 [53.3%] female; mean age 76.5 *±* 10.2 years). The training set included 29.4% positive cases, while the test set contained 3.5% positive cases.

#### 4.2.1. Result for Cognitive Decline Detection

The cognitive decline detection results of the proposed CD-Tron model and several other competing methods are shown in Table 2.

**Table 2:**
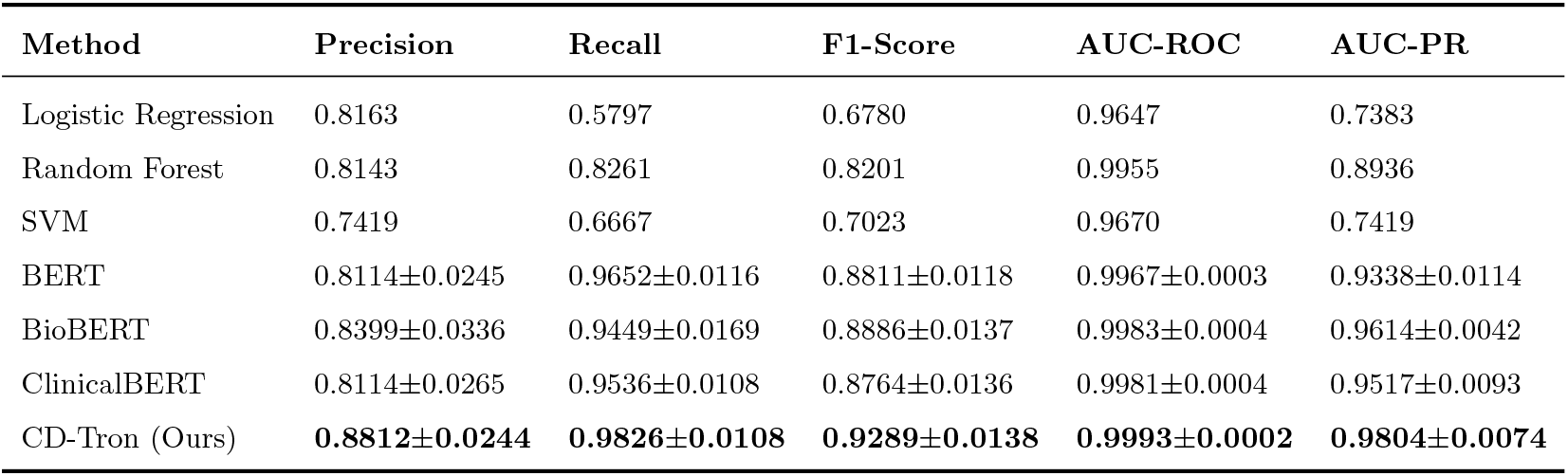
Performance of Various Methods in the Task of Cognitive Decline Detection.

| Method | Precision | Recall | F1-Score | AUC-ROC | AUC-PR |
| --- | --- | --- | --- | --- | --- |
| Logistic Regression | 0.8163 | 0.5797 | 0.6780 | 0.9647 | 0.7383 |
| Random Forest | 0.8143 | 0.8261 | 0.8201 | 0.9955 | 0.8936 |
| SVM | 0.7419 | 0.6667 | 0.7023 | 0.9670 | 0.7419 |
| BERT | 0.8114±0.0245 | 0.9652±0.0116 | 0.8811±0.0118 | 0.9967±0.0003 | 0.9338±0.0114 |
| BioBERT | 0.8399±0.0336 | 0.9449±0.0169 | 0.8886±0.0137 | 0.9983±0.0004 | 0.9614±0.0042 |
| ClinicalBERT | 0.8114±0.0265 | 0.9536±0.0108 | 0.8764±0.0136 | 0.9981±0.0004 | 0.9517±0.0093 |
| CD-Tron (Ours) | **0.8812±0.0244** | **0.9826±0.0108** | **0.9289±0.0138** | **0.9993±0.0002** | **0.9804±0.0074** |

We also plot the ROC curves and Precision-Recall curves of our CD-Tron and other models in the task of CD detection, as shown in Fig. 3.

**Figure 3:**
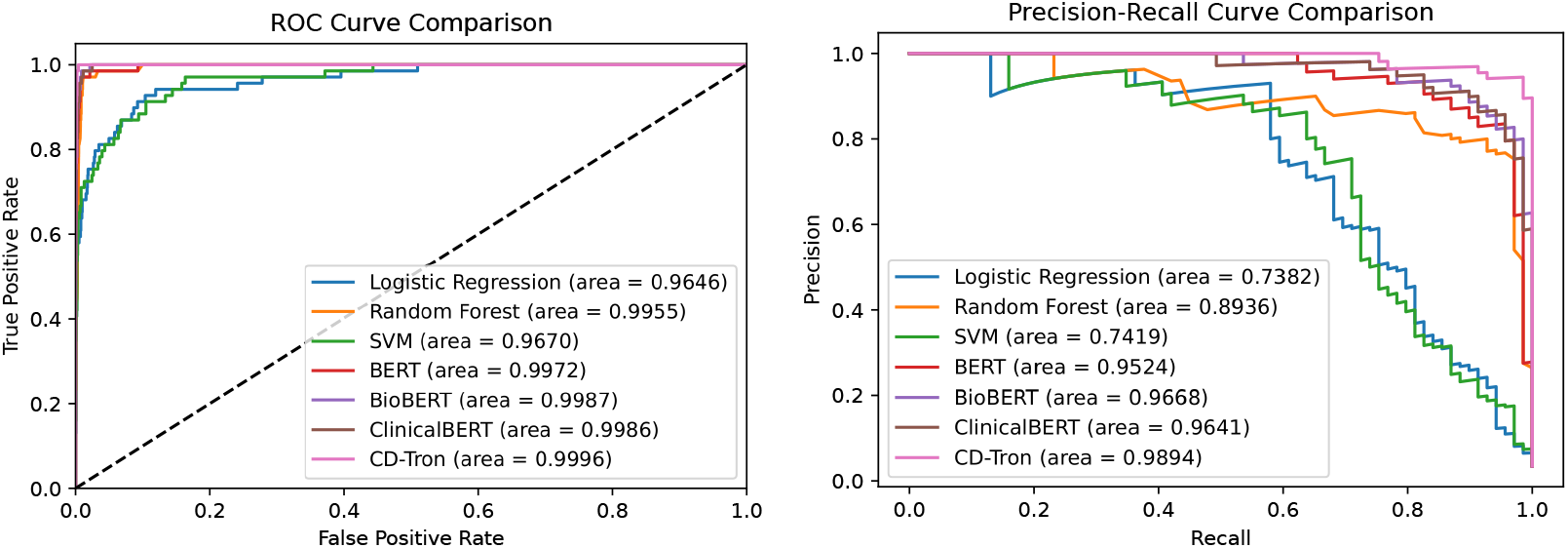
Comparison of ROC curve and precision-recall curve of different methods in the task of CD identification.

From the experimental results, we have the following observations.

- Compared with the conventional features engineering (*i.e*., TF-IDF features), the pre-trained language models achieved significantly higher performance. This demonstrated the superior feature learning capabilities of deep learning-based models, particularly in capturing complex patterns from clinical notes data.
- The pre-trained language models achieved notably higher recall compared to other methods, demonstrating their ability to effectively identify progressive CD cases. This is particularly valuable in clinical applications, where it is crucial to prioritize the comprehensive detection of patients at risk for Alzheimer’s disease, even at the expense of some false positives.
- The proposed CD-Tron model outperformed other pre-trained language models across all evaluation metrics. This can be attributed to CD-Tron’s pre-training on a significantly larger corpus of clinical notes compared to the other models, which were trained on plain texts, medical articles, or smaller datasets of clinical notes. These results underscored the advantage of large-scale clinical language models in effectively processing unstructured EHR data.

#### 4.2.2. Comparison with State-Of-The-Art LLM

We also compare the performance of the proposed CD-Tron with a state-of-the-art LLM. Specifically, GPT-4, a general-purpose LLM, was used for comparison. GPT-4 was applied to the CD detection task using prompt-based learning, where structured prompts (including required task descriptions and optional sections for prompt augmentation) were designed to guide the model in identifying cognitive decline in clinical notes.

GPT-4 achieved a precision of 71.7%, a recall of 91.4%, and an F1 score of 80.3%. In contrast, CD-Tron significantly outperformed GPT-4, achieving a precision of 88.1%, a recall of 98.3%, and an F1 score of 92.9%. This substantial performance gap highlights the value of CD-Tron as a domain-specific clinical LLM, demonstrating that incorporating clinical knowledge leads to significant improvements over general-purpose state-of-the-art LLMs.

#### 4.2.3. Explaining Model Predictions Using SHAP Values

To better understand the decision-making process of the proposed CD-Tron model, we utilized SHAP values to interpret its predictions. SHAP values provided a visual breakdown of how each token or word in the clinical notes contributes to the models decision, helping to explain the predictions for progressive CD cases.

As shown in Fig. 4, we visualized the SHAP values and highlighted the relevant text for two individual sections that were correctly classified as positive samples (*i.e*., CD) by CD-Tron. The result revealed that certain texts related to memory loss strongly influenced the model’s prediction, demonstrating how the model identifies key clinical features in the notes that were indicative of cognitive decline.

**Figure 4:**
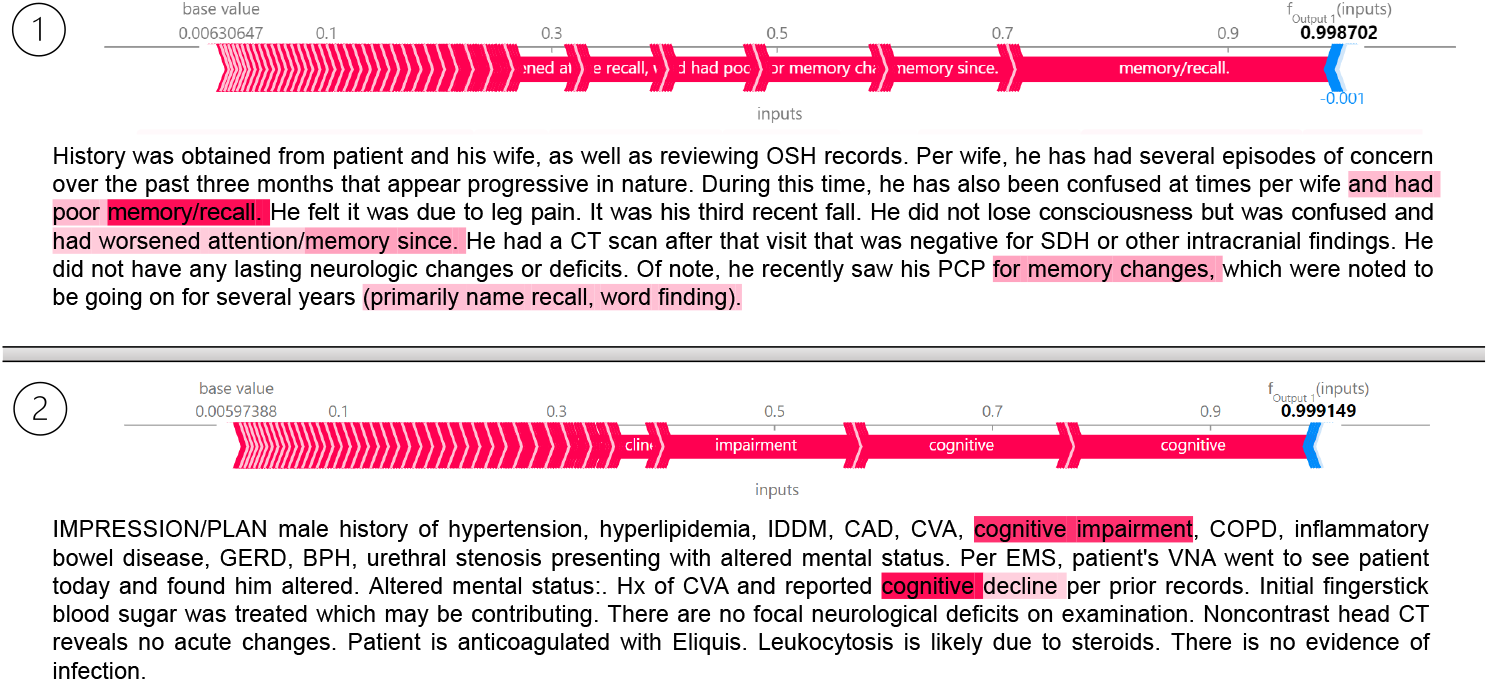
SHAP values with highlighted tokens in two individual sections that have been correctly classified as positive samples (*i.e*., CD) by the proposed CD-Tron model.

#### 4.2.4. Key Words that Influence Model Prediction

In addition to analyzing individual predictions, SHAP values can be used to identify common key words across the entire test set that significantly influence the model’s predictions for progressive CD cases. By aggregating SHAP values for all identified positive samples, we can generate an overall view of which terms contribute most strongly to the model’s decision-making process.

For this analysis, we computed the SHAP values for all positive samples in the test set and average them to quantify the overall contribution of different tokens. This process identified the most influential words that consistently appeared in the clinical notes related to CD predictions. As visualized in Fig. 5, terms related to memory, cognition, and behavioral changes (such as language fluency, which reflects difficulties in word retrieval and speech production) emerged as key factors driving the model’s predictions. This aggregated explanation provided clinicians with valuable insight into the underlying patterns the model detects in clinical notes.

**Figure 5:**
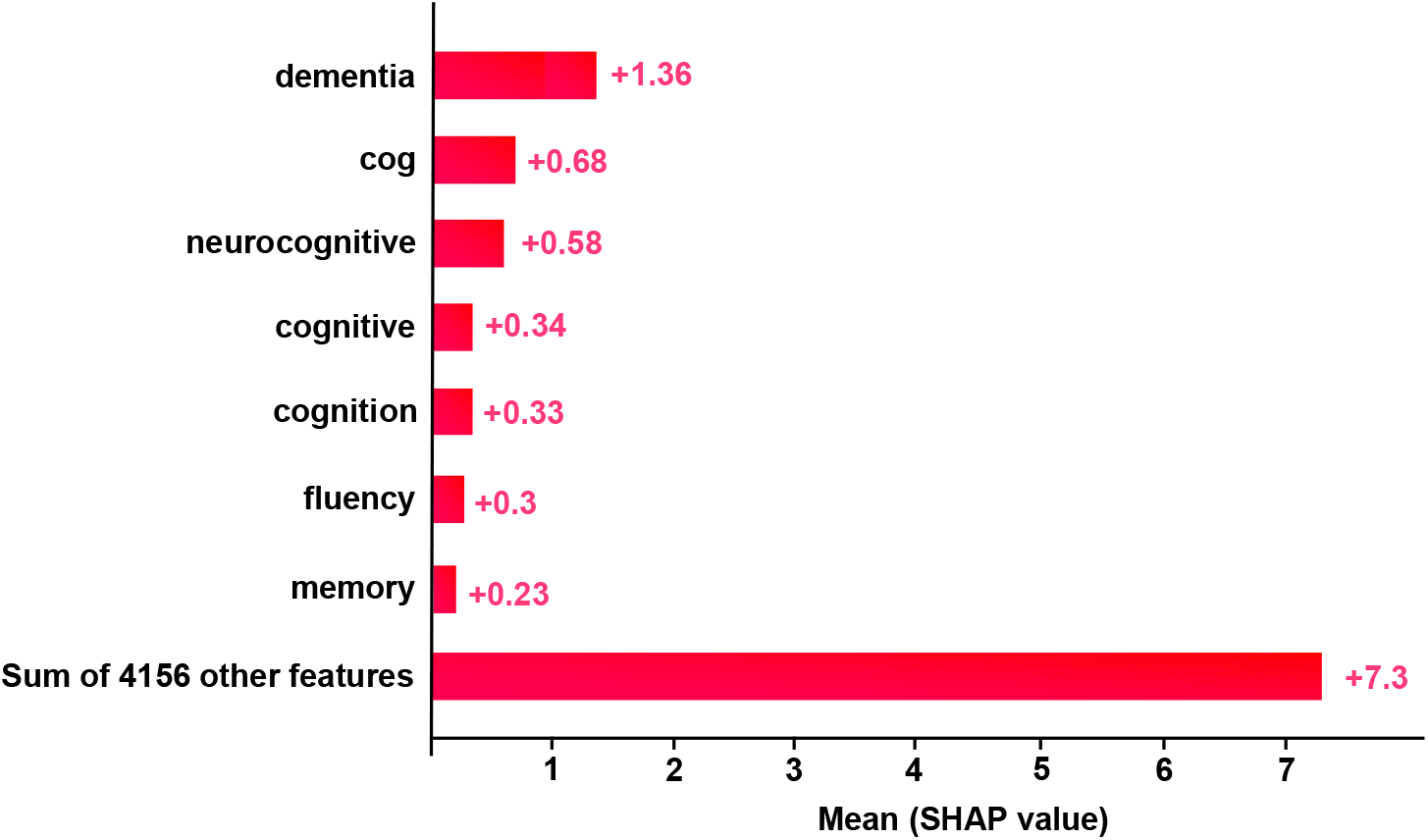
Important features for CD identification in terms of SHAP values selected by the proposed CD-Tron model.

Note that templated text like cognitive assessments which may mention terms like memory, may appear in clinical notes. However, this has minimal influence on our method due to the following measures. 1) We carefully preprocessed the dataset to minimize the impact of templated sections. This included manual reviews of sample notes to avoid templated terms. 2) SHAPs ability to evaluate the contextual contribution of each feature within the **entire input** ensures that frequently occurring templated text without meaningful relevance to the model’s predictions receives low SHAP values. This guarantees that such terms do not significantly influence the model’s decisions.

#### 4.2.5. Error Analysis

To further understand the prediction of the proposed CD-Tron model, we conducted error analysis by exploring the false positive and false negative prediction cases by the model.

We first visualized the confusion matrix of the model’s prediction result, as shown in Fig. 6. From the results, it is evident that the model achieves high sensitivity, with only one false negative case. This is crucial in clinical practice, as missing positive cases can have more severe consequences than false alarms. On the other hand, there are 8 false positive predictions.

**Figure 6:**
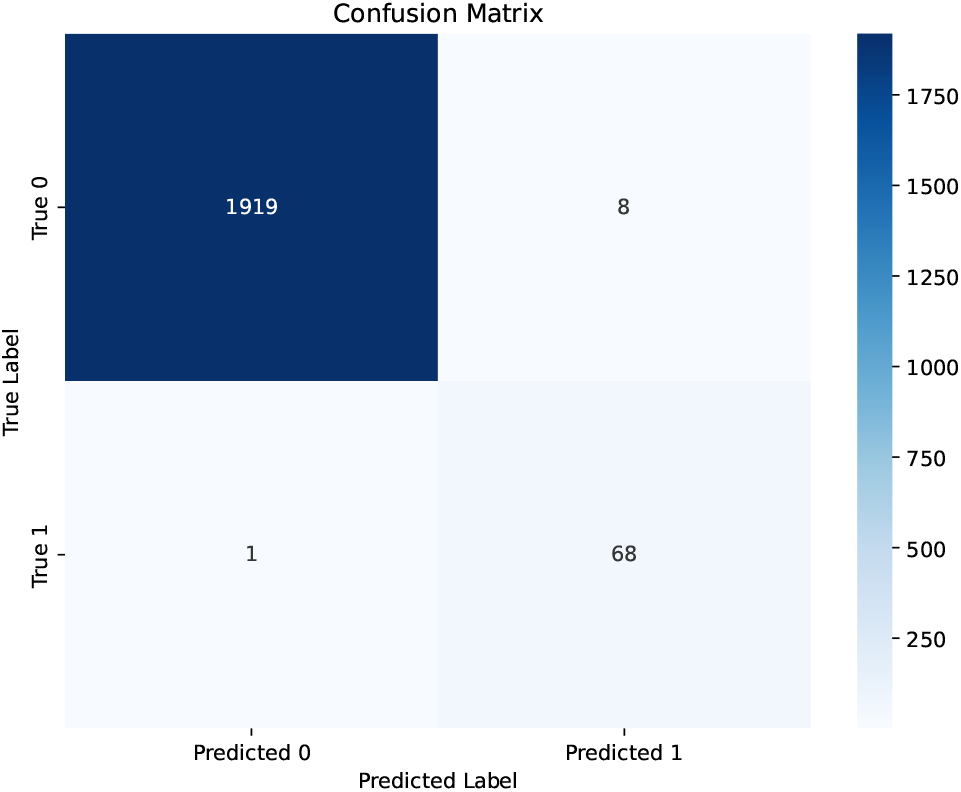
Confusion matrix of the model’s prediction. Label 1 represents CD while label 0 denotes non-CD.

Table 3 presents the representative note sections corresponding to both the false negative and false positive cases, along with explanations for the errors made by the model.

**Table 3:** Error Analysis for False Negative/Positive Predictions of the CD-Tron Model.

| Potential Reasons | Representative Cases |
| --- | --- |
| Weak Evidence. Primarily focuses on physical impairments and care goals, with minimal mention of cognitive symptoms, potentially leading to misclassification | **False Negative.**<br>GOALS HH - ADL / IADL Impairment. HH - Promote higher level of independence with performance of ADLs/IADLs. [OT]. HH - Advance Care Planning. HH - Verbalize awareness of Advance Care Planning. [MSW, OT, PT, SLP, SN]. HH - Aspiration - Risk of. HH - Knowledge and management of aspiration precautions. [MSW, OT, PT, SLP, SN]. ... HH - Health Maintenance / Patient Strengths Impacting Goal Achievement. HH - Patient will utilize their strengths to achieve optimal knowledge and management for safe home/ community functioning. [MSW, OT, PT, SLP, SN]. HH - Mental Status - Impaired. HH - Mental status allows patient to safely function within their home environment... |
| The issue is caused by side effects of medication rather than CD. | **False Positive.**<br>Onc.: Clinically doing well on Ibrutinib. However the patient and her husband are concerned that her memory and anxiety/depression has worsened on this medication. We discussed this with Dr. A and Dr. B as well. We offered holding the ibrutinib to see if this would help. However she is not interested in doing this at this time. We discussed other options such as Venetoclax. She prefers to treat the depression instead and stay on Ibrutinib. |

## 5. Discussion

The findings of this study demonstrate the potential of the CD-Tron model to address a critical gap in clinical practice: the early detection of cognitive decline using clinical notes. By leveraging a large clinical language model, CD-Tron not only advances the processing of electronic health records (EHRs) but also provides valuable insights that can directly impact clinical workflows and patient care.

### 5.1. Clinical Applications and Benefits

The CD-Tron model offers clinicians a valuable tool for identifying patients at risk of CD, a precursor to AD. By automating the analysis of clinical notes, the model can reduce manual effort, integrates seamlessly into existing clinical workflows, and enables efficient prioritization of at-risk patients for further testing or specialist referrals. Moreover, its ability to enhance diagnostic accuracy supports earlier interventions, streamlining clinical decision-making. These capabilities can ultimately improve patient outcomes and optimize healthcare resource utilization by minimizing the burden on clinicians.

### 5.2. Broader Implications for Patient Care

The CD-Tron model showcases a broader impact on early disease screening and patient care through the use of clinical notes. Its proactive approach to detecting cognitive decline supports timely interventions and care, potentially slowing or delaying progression to AD and improving the quality of life for patients and their families. Furthermore, this methodology holds promise for application in the early screening of other diseases using unstructured EHR data.

### 5.3. Contextualizing Findings Within Existing Research

This study builds on prior research that applies machine learning and natural language processing techniques to CD detection. Traditional methods often rely on structured data or are limited in their ability to process the rich information embedded in free-text clinical notes. By leveraging GatorTron, a large clinical language model pre-trained on extensive clinical data, CD-Tron addresses these limitations, offering enhanced feature extraction and a more nuanced understanding of patient records. This capability underscores the importance of using domain-specific language models to tackle complex medical challenges.

### 5.4. Limitations

While CD-Tron demonstrated promising results in detecting early cognitive decline, several limitations should be acknowledged. First, our study was based on a specific dataset from a single healthcare system, which may limit the model’s applicability to other populations from other healthcare systems. Additionally, since the model is designed to detect CD based on cognitive complaints explicitly documented in the notes, it might not identify those patients who progress directly to MCI or AD without having any prior subjective cognitive complaints recorded, either because such complaints were not expressed or were not documented by clinicians.

Future work will focus on expanding the scope of CD-Tron to incorporate multi-modal data, such as MRI scans or genetic information, to improve detection accuracy. Additionally, we plan to extend the model’s validation to broader and more diverse patient populations across different healthcare systems to ensure its robustness and generalizability.

## 6. Conclusion

In this paper, we present CD-Tron, a novel approach for the early detection of cognitive decline using clinical notes from real-world EHRs. By leveraging the power of large-scale clinical language models, CD-Tron is designed to detect early signs of CD, a critical precursor to AD/ADRD. Our approach adapts state-of-the-art clinical language models to handle the complexities of unstructured clinical data, providing a robust tool for early diagnosis and intervention. Additionally, the integration of SHAP for model interpretability enhances the transparency of predictions, offering clinicians valuable insights into the factors influencing the models decisions. This combination of advanced modeling and explainability makes CD-Tron a trustworthy and impactful tool for clinical practice.

## Data Availability

The data used in this study are not publicly available due to institutional and patient privacy restrictions.

## Acknowledgments

This study was funded by NIH-NIA R01AG080429.

## References

[1] C. L. Masters, R. Bateman, K. Blennow, C. C. Rowe, R. A. Sperling, J. L. Cummings, Alzheimer’s disease, Nature Reviews Disease Primers 1 (1) (2015) 1–18.

[2] F. B. Ahmad, R. N. Anderson, The leading causes of death in the US for 2020, JAMA 325 (18) (2021) 1829–1830.

[3] E. A. Kramarow, B. Tejada-Vera, National vital statistics reports, National Vital Statistics Reports 68 (2) (2019) 1–18.

[4] A. Nandi, N. Counts, J. Bröker, S. Malik, S. Chen, R. Han, J. Klusty, B. Seligman, D. Tortorice, D. Vigo, et al., Cost of care for Alzheimers disease and related dementias in the United States: 2016 to 2060, NPJ Aging 10 (1) (2024) 1–8.

[5] D. W. Scharre, Preclinical, prodromal, and dementia stages of Alzheimer’s disease, Practical Neurology 15 (2019) 36–47.

[6] L. A. Rabin, C. M. Smart, R. E. Amariglio, Subjective cognitive decline in preclinical Alzheimer’s disease, Annual review of clinical psychology 13 (1) (2017) 369–396.

[7] M. Crous-Bou, C. Minguillón, N. Gramunt, J. L. Molinuevo, Alzheimers disease prevention: from risk factors to early intervention, Alzheimer’s Research & Therapy 9 (2017) 1–9.

[8] L. Robinson, E. Tang, J.-P. Taylor, Dementia: timely diagnosis and early intervention, BMJ 350 (2015).

[9] P. Arrondo, Ó. Elía-Zudaire, G. Martí-Andrés, M. A. Fernández-Seara, M. Riverol, Grey matter changes on brain MRI in subjective cognitive decline: A systematic review, Alzheimer’s Research & Therapy 14 (1) (2022) 1–16.

[10] J. Lagarde, P. Olivieri, M. Tonietto, C. Tissot, I. Rivals, P. Gervais, F. Caillé, M. Moussion, M. Bottlaender, M. Sarazin, Tau-PET imaging predicts cognitive decline and brain atrophy progression in early Alzheimer’s disease, Journal of Neurology, Neurosurgery & Psychiatry 93 (5) (2022) 459–467.

[11] S. Wolfsgruber, A. Polcher, A. Koppara, L. Kleineidam, L. Frölich, O. Peters, M. Hüll, E. Rüther, J. Wiltfang, W. Maier, et al., Cerebrospinal fluid biomarkers and clinical progression in patients with subjective cognitive decline and mild cognitive impairment, Journal of Alzheimer’s Disease 58 (3) (2017) 939–950.

[12] R. Sherva, A. Gross, S. Mukherjee, R. Koesterer, P. Amouyel, C. Bellenguez, C. Dufouil, D. A. Bennett, L. Chibnik, C. Cruchaga, et al., Genome-wide association study of rate of cognitive decline in Alzheimer’s disease patients identifies novel genes and pathways, Alzheimer’s & Dementia 16 (8) (2020) 1134–1145.

[13] Q. Li, X. Yang, J. Xu, Y. Guo, X. He, H. Hu, T. Lyu, D. Marra, A. Miller, G. Smith, et al., Early prediction of Alzheimer’s disease and related dementias using real-world electronic health records, Alzheimer’s & Dementia 19 (8) (2023) 3506–3518.

[14] P. Yadav, M. Steinbach, V. Kumar, G. Simon, Mining electronic health records (EHRs): A survey, ACM Computing Surveys (CSUR) 50 (6) (2018) 1–40.

[15] E. Hossain, R. Rana, N. Higgins, J. Soar, P. D. Barua, A. R. Pisani, K. Turner, Natural language processing in electronic health records in relation to healthcare decision-making: A systematic review, Computers in Biology and Medicine 155 (2023) 1–24.

[16] L. Wang, J. Laurentiev, J. Yang, Y.-C. Lo, R. E. Amariglio, D. Blacker, R. A. Sperling, G. A. Marshall, L. Zhou, Development and validation of a deep learning model for earlier detection of cognitive decline from clinical notes in electronic health records, JAMA Network Open 4 (11) (2021) 1–11.

[17] D. Khurana, A. Koli, K. Khatter, S. Singh, Natural language processing: state of the art, current trends and challenges, Multimedia Tools and Applications 82 (3) (2023) 3713–3744.

[18] I. Li, J. Pan, J. Goldwasser, N. Verma, W. P. Wong, M. Y. Nuzumlalı, B. Rosand, Y. Li, M. Zhang, D. Chang, et al., Neural natural language processing for unstructured data in electronic health records: a review, Computer Science Review 46 (2022) 1–29.

[19] J. Devlin, M.-W. Chang, K. Lee, K. Toutanova, BERT: Pre-training of deep bidirectional transformers for language understanding, in: Proceedings of the 2019 Conference of the North American Chapter of the Association for Computational Linguistics: Human Language Technologies), Association for Computational Linguistics, 2019, pp. 4171–4186.

[20] K. Huang, J. Altosaar, R. Ranganath, ClinicalBERT: Modeling clinical notes and predicting hospital readmission, arXiv preprint arXiv:1904.05342 (2019).

[21] J. Lee, W. Yoon, S. Kim, D. Kim, S. Kim, C. H. So, J. Kang, BioBERT: a pre-trained biomedical language representation model for biomedical text mining, Bioinformatics 36 (4) (2020) 1234–1240.

[22] L. Rasmy, Y. Xiang, Z. Xie, C. Tao, D. Zhi, Med-BERT: pretrained contextualized embeddings on large-scale structured electronic health records for disease prediction, NPJ Digital Medicine 4 (1) (2021) 1–13.

[23] B. Wang, Q. Xie, J. Pei, Z. Chen, P. Tiwari, Z. Li, J. Fu, Pre-trained language models in biomedical domain: A systematic survey, ACM Computing Surveys 56 (3) (2023) 1–52.

[24] P. Hager, F. Jungmann, R. Holland, K. Bhagat, I. Hubrecht, M. Knauer, J. Vielhauer, M. Makowski, R. Braren, G. Kaissis, et al., Evaluation and mitigation of the limitations of large language models in clinical decision-making, Nature Medicine (2024) 1–10.

[25] K. S. Kalyan, A. Rajasekharan, S. Sangeetha, Ammu: a survey of transformer-based biomedical pretrained language models, Journal of Biomedical Informatics 126 (2022) 1–23.

[26] C. Mao, J. Xu, L. Rasmussen, Y. Li, P. Adekkanattu, J. Pacheco, B. Bonakdarpour, R. Vassar, L. Shen, G. Jiang, et al., AD-BERT: Using pre-trained language model to predict the progression from mild cognitive impairment to Alzheimer’s disease, Journal of Biomedical Informatics 144 (2023) 1–8.

[27] I. Y. Oh, S. E. Schindler, N. Ghoshal, A. M. Lai, P. R. Payne, A. Gupta, Extraction of clinical phenotypes for Alzheimer’s disease dementia from clinical notes using natural language processing, JAMIA Open 6 (1) (2023) 1–9.

[28] L. C. Maclagan, M. Abdalla, D. A. Harris, T. A. Stukel, B. Chen, E. Candido, R. H. Swartz, A. Iaboni, R. L. Jaakkimainen, S. E. Bronskill, Can patients with dementia be identified in primary care electronic medical records using natural language processing?, Journal of Healthcare Informatics Research 7 (1) (2023) 42–58.

[29] W. X. Zhao, K. Zhou, J. Li, T. Tang, X. Wang, Y. Hou, Y. Min, B. Zhang, J. Zhang, Z. Dong, et al., A survey of large language models, arXiv preprint arXiv:2303.18223 (2023).

[30] S. Minaee, T. Mikolov, N. Nikzad, M. Chenaghlu, R. Socher, X. Amatriain, J. Gao, Large language models: A survey, arXiv preprint arXiv:2402.06196 (2024).

[31] H. Naveed, A. U. Khan, S. Qiu, M. Saqib, S. Anwar, M. Usman, N. Akhtar, N. Barnes, A. Mian, A comprehensive overview of large language models, arXiv preprint arXiv:2307.06435 (2023).

[32] A. J. Thirunavukarasu, D. S. J. Ting, K. Elangovan, L. Gutierrez, T. F. Tan, D. S. W. Ting, Large language models in medicine, Nature Medicine 29 (8) (2023) 1930–1940.

[33] K. Singhal, S. Azizi, T. Tu, S. S. Mahdavi, J. Wei, H. W. Chung, N. Scales, A. Tanwani, H. Cole-Lewis, S. Pfohl, et al., Large language models encode clinical knowledge, Nature 620 (7972) (2023) 172–180.

[34] Z. A. Nazi, W. Peng, Large language models in healthcare and medical domain: A review, in: Informatics, Vol. 11, MDPI, 2024, pp. 1–23.

[35] X. Du, J. Novoa-Laurentiev, J. M. Plasek, Y.-W. Chuang, L. Wang, G. A. Marshall, S. K. Mueller, F. Chang, S. Datta, H. Paek, et al., Enhancing early detection of cognitive decline in the elderly: a comparative study utilizing large language models in clinical notes, EBioMedicine 109 (2024).

[36] X. Yang, A. Chen, N. PourNejatian, H. C. Shin, K. E. Smith, C. Parisien, C. Compas, C. Martin, A. B. Costa, M. G. Flores, et al., A large language model for electronic health records, NPJ digital medicine 5 (1) (2022) 1–9.

[37] L. Y. Jiang, X. C. Liu, N. P. Nejatian, M. Nasir-Moin, D. Wang, A. Abidin, K. Eaton, H. A. Riina, I. Laufer, P. Punjabi, et al., Health system-scale language models are all-purpose prediction engines, Nature 619 (7969) (2023) 357–362.

[38] F. Jessen, R. E. Amariglio, M. Van Boxtel, M. Breteler, M. Ceccaldi, G. Chételat, B. Dubois, C. Dufouil, K. A. Ellis, W. M. Van Der Flier, et al., A conceptual framework for research on subjective cognitive decline in preclinical Alzheimer’s disease, Alzheimer’s & Dementia 10 (6) (2014) 844–852.

[39] L. Zhou, J. M. Plasek, L. M. Mahoney, N. Karipineni, F. Chang, X. Yan, F. Chang, D. Dimaggio, D. S. Goldman, R. A. Rocha, Using medical text extraction, reasoning and mapping system (MTERMS) to process medication information in outpatient clinical notes, in: AMIA Annual Symposium Proceedings, Vol. 2011, American Medical Informatics Association, 2011, pp. 1639–1648.

[40] A. Vaswani, et al., Attention is all you need, Advances in Neural Information Processing Systems (2017) 1–11.

[41] S. M. Jain, Hugging face, in: Introduction to transformers for NLP: With the hugging face library and models to solve problems, Springer, 2022, pp. 51–67.

[42] M. Scott, L. Su-In, et al., A unified approach to interpreting model predictions, Advances in Neural Information Processing Systems 30 (2017) 4765–4774.

[43] R. Dwivedi, D. Dave, H. Naik, S. Singhal, R. Omer, P. Patel, B. Qian, Z. Wen, T. Shah, G. Morgan, et al., Explainable AI (XAI): Core ideas, techniques, and solutions, ACM Computing Surveys 55 (9) (2023) 1–33.

[44] S. Jain, B. C. Wallace, Attention is not explanation, in: Proceedings of the 2019 Conference of the North American Chapter of the Association for Computational Linguistics: Human Language Technologies, Volume 1 (Long and Short Papers), 2019, pp. 3543–3556.

[45] S. Serrano, N. A. Smith, Is attention interpretable?, in: Proceedings of the 57th Annual Meeting of the Association for Computational Linguistics, 2019, pp. 2931–2951.

